# Perceived value and stakeholder experience of a dedicated allied health professions informatics role in a specialist cancer centre: a mixed-methods service evaluation survey

**DOI:** 10.64898/2026.09.11.26362711

**Authors:** Derek Dale Bayquen

**Affiliations:** The Royal Marsden NHS Foundation Trust, London, UK; University of Winchester, Winchester, UK

## Abstract

**Background:** Dedicated allied health professions (AHP) informatics posts remain rare and largely unevaluated: one NHS trust in ten reports a formal AHP informatics lead.

**Objectives:** To evaluate stakeholder-perceived value and experience of a dedicated AHP Information Officer (AHPIO) role supporting an organisation-wide electronic health record (EHR), and to derive a working framework of the role’s practice.

**Methods:** Cross-sectional online survey within a registered service evaluation at a specialist cancer centre in England. A purposive census of 40 stakeholders (clinical and operational AHP staff, digital champions and EHR programme colleagues) rated six agreement items with four free-text questions. Analysis led with distributions, medians and Wilson intervals; inference was exploratory; free-text coding is fully audit-trailed.

**Results:** Twenty-seven stakeholders responded (68%). Five items sat at a ceiling (medians 4 to 5; agreement 95% to 100%); all 25 answering the overall item agreed the role adds value, 80% strongly. Data and reporting access was the outlier (median 3.0; 52% neutral or below; agreement 48%, 95% CI 28% to 68%), ranking below every other item (Friedman chi-squared 36.3, df 5, 20 complete cases; Holm-adjusted post-hoc comparisons, rank-biserial -0.73 to -1.00). Nine themes located value in brokerage and routing (18/26), concrete outcome accounts (16/26) and a clinician who understands AHP work (12/26).

**Conclusions:** Stakeholders located the role’s value in brokerage, clinical-digital translation, representation and outcome response, resting on a named trusted contact; self-service data is the part-built pillar and development priority. The framework and method offer a replicable template for evaluating AHP informatics roles.

## Introduction

Allied health professionals (AHPs) are the third largest clinical workforce in the NHS, spanning fourteen professions whose daily work runs through shared electronic health records they rarely helped to shape. What Good Looks Like, the NHS digital-maturity framework, makes a well-led, digitally capable workforce a marker of organisational maturity.^1^ Assessed against exactly that frame, the first national AHP Digital Maturity Assessment found AHP digital leadership the weakest pillar in the country at 28%, with decision support and analytics next at roughly 34% to 40%, records, governance and remote care the strongest areas, and only about one organisation in ten reporting a formal AHP informatics lead.^2^ The workforce’s own competency reference points the same way: the national AHP digital competency framework carries meta-competencies and digital leadership as a domain of practice for every profession, and its published evaluation found the framework too abstract for specific roles, with awareness of it near zero, so the leadership and meta-competency layer is simultaneously the weakest assessed and the least operationalised.^3 4^ Chief clinical and nursing information officer roles are established features of NHS digital leadership; their AHP equivalents are emergent, thinly spread and, so far, largely unevaluated.

The case for profession-specific informatics capacity rests on a simple mechanism: EHR value depends on continuing, clinically informed refinement after go-live, and that refinement depends on people who hold both the clinical workflow and the system.^5 6^ Where such roles exist for other professions they concentrate on exactly this seam. What is missing is evidence about what a dedicated AHP post actually does for its stakeholders, and how the people who deal with it judge its value. Much of the work such a role performs is demand-modifying and brokered: it prevents problems, routes questions and translates between worlds, which activity counts do not capture, so its value stays invisible unless stakeholders are asked directly.

This study evaluates a dedicated AHP Information Officer (AHPIO) post at a specialist cancer centre in England, about six months after the post was created, on an organisation-wide EHR live for three years. It asks three questions. How do stakeholders rate the support across its intended functions? What do they say the role changes, in their own words? And what working framework of AHPIO practice and perceived value do the two strands together suggest, as a template other organisations could test?

## Methods

### Design and setting

A cross-sectional online stakeholder survey conducted within a registered service evaluation (amendment) under the Health Research Authority decision framework^11^ at a specialist cancer centre in England running a single organisation-wide EHR across all clinical services. Reporting follows the Checklist for Reporting Results of Internet E-Surveys (CHERRIES; Supplementary File, section S4).^8^

### THE role

The AHPIO is a single whole-time clinically qualified post created to support AHP digital practice: routing and escalating EHR issues, coordinating requests for system improvement, running walkarounds and a digital champion structure, liaising with training, and representing AHP requirements in digital governance. The EHR had run for three years without the post; the evaluation therefore asks what the dedicated role adds to a functioning system.

### Participants

A purposive census of the post’s 40 known stakeholders was invited by email: operational and professional leads across AHP and adjacent services, digital champions and superusers, administrative leads, and EHR programme colleagues (analysts and trainers) who work with the post. The programme group co-produces some of the work it rates, and this non-independence is declared and handled in analysis. No a priori sample size calculation was performed; the frame is a census and inference is exploratory throughout. The stakeholder list was enumerated from the post’s working registers: the professional and operational lead directory, the digital champion roster and the EHR programme’s team list, so the frame is a census of known stakeholders rather than a sample of a wider population. No incentive was offered.

### Instrument

Six five-point agreement items (strongly disagree to strongly agree) were written in-house from the themes of a local AHP digital-practice inventory, covering knowing who to approach, acknowledgement and escalation of raised issues, perceived improvement or safety of the service’s EHR setup, perceived action on digital risk, access to data, reports and dashboards, and overall added value. Four free-text prompts asked for context, a specific occasion when the support made a difference or was missing, what works and what does not, and what would change if the post were removed. Profile items recorded role, service, site and support frequency. The form was checked with one internal test submission before launch, which was removed from the dataset. The instrument is reproduced verbatim in the Supplementary File (S1). Two items (setup improvement and risk response) record perceptions of outcomes, deliberately, and are reported as perceptions throughout.

### Data collection

The survey ran as a closed online form from 26 May to 16 June 2026. Responses were non-anonymous by design (a named-contact field was optional and half completed it), because the survey rates named support; ceiling-inflated positivity is therefore expected and is handled in interpretation. Participation was voluntary and submission was taken as consent. Responses were held on the organisation’s survey platform, and names and email addresses were removed before analysis. Twenty-eight records were returned; one 73-second test record was removed, leaving an analytic sample of 27.

### Analysis

The items are ordinal, so analysis leads with response distributions, medians and top-box percentages, with means and standard deviations reported for convenience. Wilson 95% intervals bound the headline proportions.^10^ Inferential tests are exploratory because the sample is purposive and self-selected: a within-respondent Friedman test across the six items, with post-hoc Wilcoxon signed-rank comparisons of the data-access item against each of the other five items (a many-to-one family selected after inspection of the distributions, Holm-adjusted within that family and read descriptively) with rank-biserial effect sizes, one-sample Wilcoxon tests against the scale midpoint, and Mann-Whitney comparisons between the clinical and programme groups. Internal consistency is reported as indicative only. Free-text responses were coded by the author, the sole researcher, into nine themes,^9^ with an analytic memo recording the coding decisions, the rules applied and the alternative readings considered (Supplementary File, S6); the full respondent-by-theme matrix is reproduced as an audit trail (S3), every count traces to it, and the complete de-identified free text is published (S5) so readers can audit or re-code every assignment. All figures were computed directly from the raw export by a build script and re-derived independently without discrepancy.

Missing responses were reported per item and not imputed; every percentage uses that item’s answered denominator, shown throughout. No weighting was applied, because the frame is a purposive census rather than a probability sample, and generalisation is not claimed. The exploratory label is load-bearing: the p-values order evidence within this sample and are not population inferences, which is why the distributions, medians and intervals lead every reading.

### Reflexivity and competing interests

The post evaluated is held by the author. Five mitigations apply: an analytic memo records the coding decisions and the alternative readings considered (Supplementary File, S6); every respondent account is reproduced verbatim (S5); the full respondent-by-theme matrix is published (S3), so readers can audit or re-code every interpretive step; every statistic is computationally reproducible from the raw export; and claims are restricted throughout to satisfaction and perceived value, never to demonstrated objective impact. Criticisms of the post are reported unsoftened.

## Results

### Respondents

Twenty-seven of the 40 invited stakeholders responded (68%): 18 clinical or operational staff (operational and professional leads, digital champions, administrative leads across occupational therapy, physiotherapy, speech and language therapy, dietetics, psychological services, lymphoedema, and diagnostic and therapeutic radiography) and nine EHR programme colleagues (analysts and trainers). Six used the support at least weekly, a further eleven at least monthly, and ten once or twice in six months (Table 1). Two respondents completed the profile and free text while skipping most rating items, which the per-item denominators in Table 2 make visible.

**Table 1.** Respondent profile (analytic N=27). Group definitions and the full service breakdown are in the Supplementary File (S2).

| Characteristic | n (of 27) |
| --- | --- |
| Clinical or operational AHP and adjacent services | 18 |
| Operational or professional lead | 10 |
| Digital champion or superuser | 5 |
| Administrative lead | 3 |
| EHR programme colleagues (analysts, trainers) | 9 |
| At least weekly | 6 |
| At least monthly, less than weekly | 11 |
| Once or twice in six months | 10 |

**Table 2.**
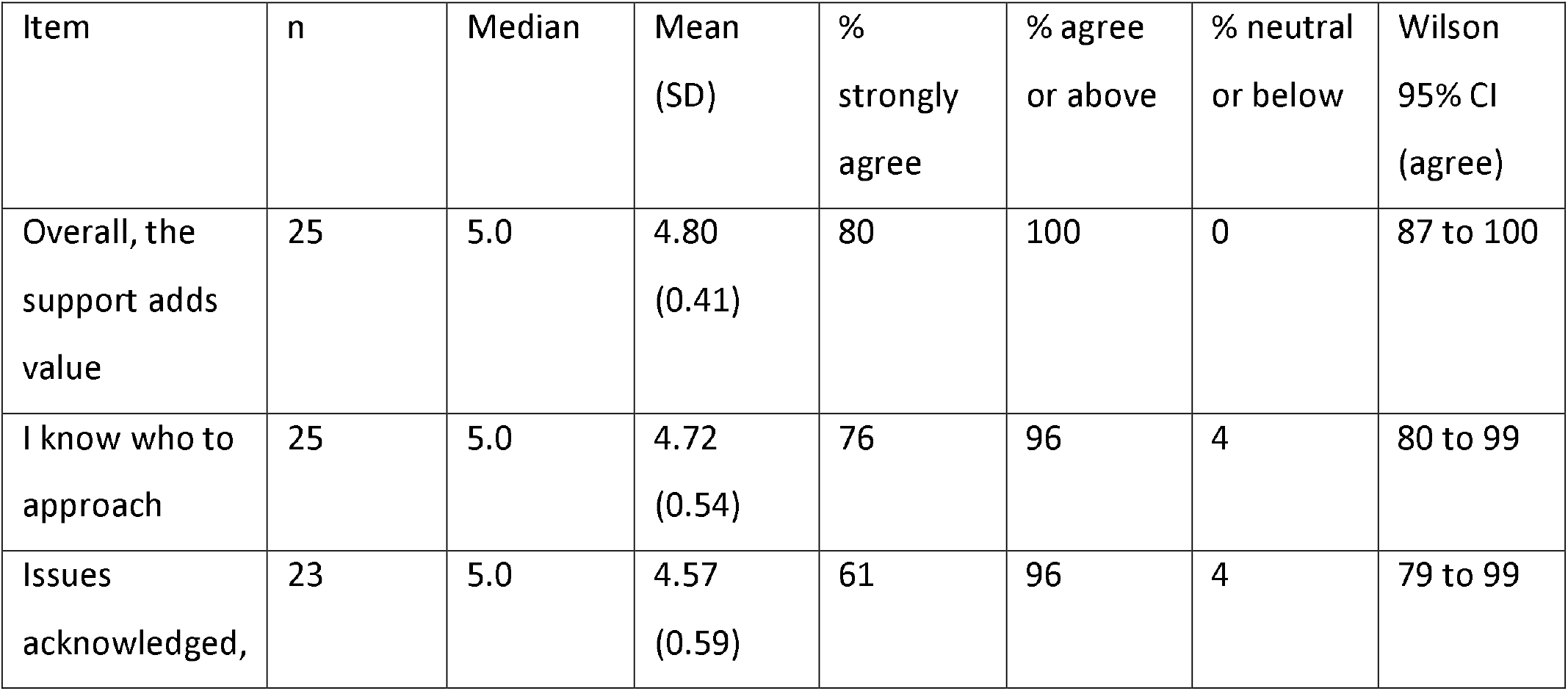
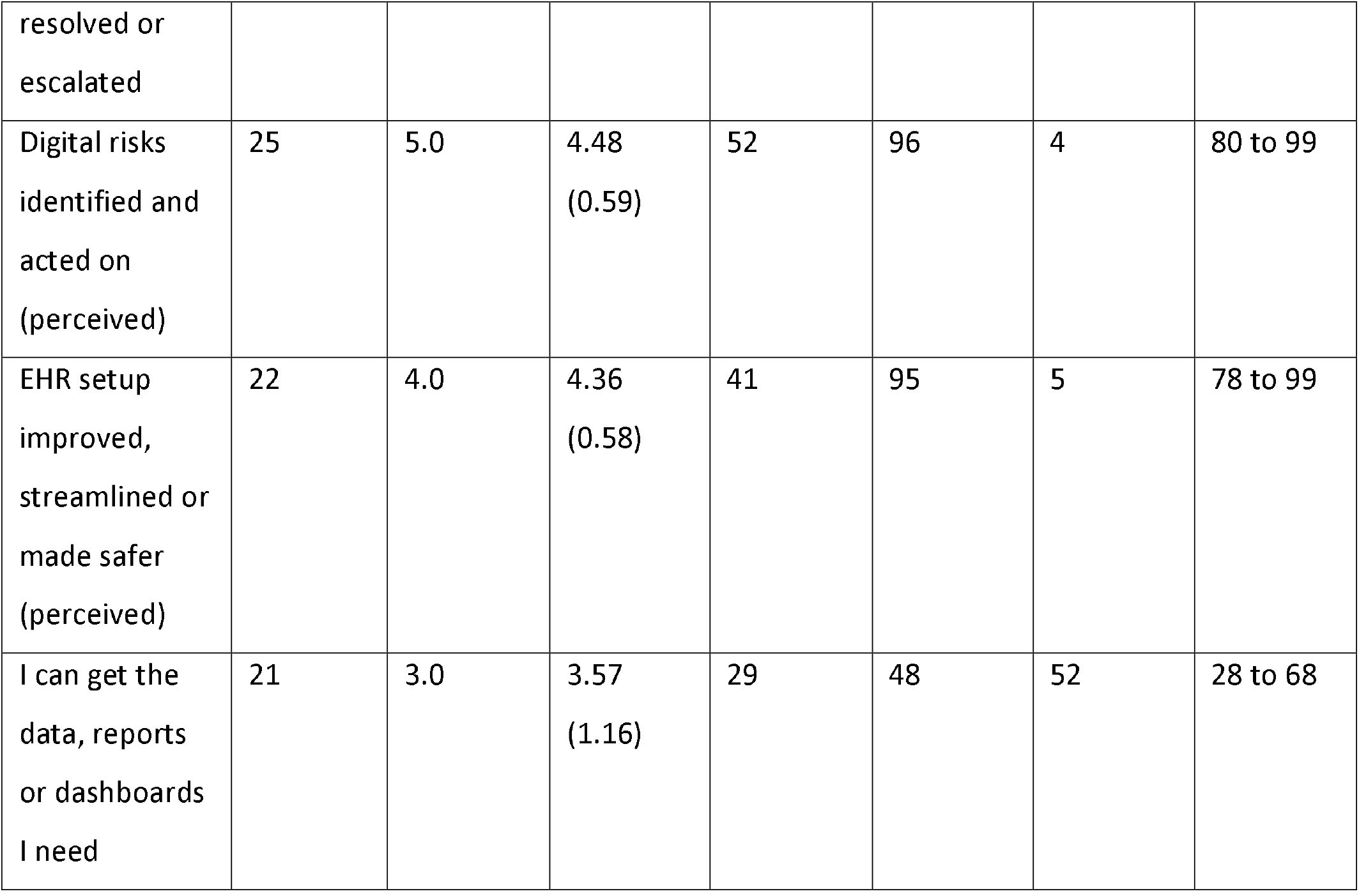
Item ratings among those answering each item (missing shown by n; two respondents skipped most rating items). Items are ordinal; medians and distributions are the primary descriptors and means are for convenience. Two items record perceptions, deliberately.

### Ratings: five ceilings and one outlier

Five of six items sat at a ceiling: medians of 4 to 5, agreement 95% to 100% among those answering, and no disagreement (Table 2; Figure 2). Every respondent who answered the overall item agreed the support adds value to the service and patient care (25 of 25), with 80% strongly agreeing; even at its Wilson lower bound the overall endorsement remains high (87%). Knowing who to approach (96% agreement) and acknowledgement and escalation of raised issues (96%) were the strongest specific functions.

Access to data, reports and dashboards was the single outlier: median 3.0 and mean 3.57 against 4.36 to 4.80 elsewhere, 52% of the 21 who answered at neutral or below, and the only item drawing any disagreement. Its agreement proportion (48%) carries a wide interval (95% CI 28% to 68%), and the shortfall is the clinical users’ own: among the 18 clinical respondents 17 answered this item, with a median of 3.0 and agreement of 41%, while most of those who skipped it were programme staff who said they were not service users of it.

The exploratory within-respondent test confirms what the distributions show: ratings differed across items (Friedman chi-squared 36.3, df 5, p<0.001, Kendall’s W 0.36, 20 complete cases), with the data-access item ranking below every other item in the Holm-adjusted five-comparison post-hoc family (rank-biserial -0.73 to -1.00) and the pattern surviving among clinical staff alone (p<0.001, 17 complete cases). One-sample tests against the midpoint restate the ceiling for five items (all p<0.001, rank-biserial +1.00) and place data access only marginally above neutral (p=0.040, rank-biserial +0.63). No clinical-versus-programme difference was detected on any item, an underpowered comparison that does not demonstrate equivalence. Six-item internal consistency was indicative only (Cronbach’s alpha 0.71; 0.83 across the four operational items).

### What stakeholders said

Twenty-six of 27 respondents left written comments, coded by the author to nine themes (Table 3). Brokerage and routing to the right digital team was the most common (18/26): “helped us coordinate with the right end users, understand the clinical requirements” (programme respondent). Concrete outcomes, efficiency, income and safety, were raised by 16 of 26 as specific episodic accounts, reported here as respondent accounts rather than verified outcomes. A clinician who understands AHP work (12/26) and a named, responsive point of contact (10/26) carried the relational core: “The escalation of our needs and the feedback is really helpful” (clinical respondent); “quick to respond to any queries” (programme respondent).

**Table 3.**
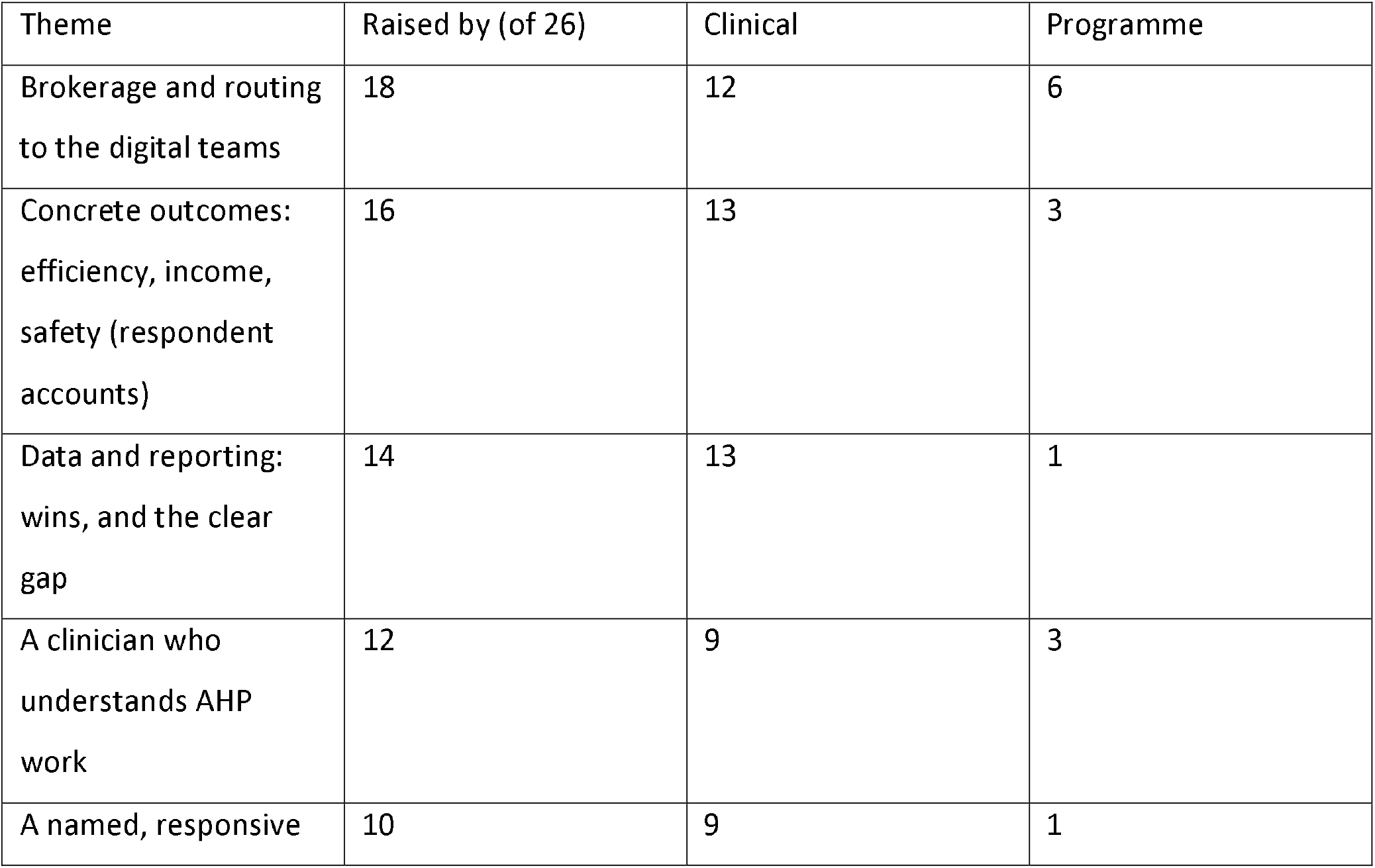
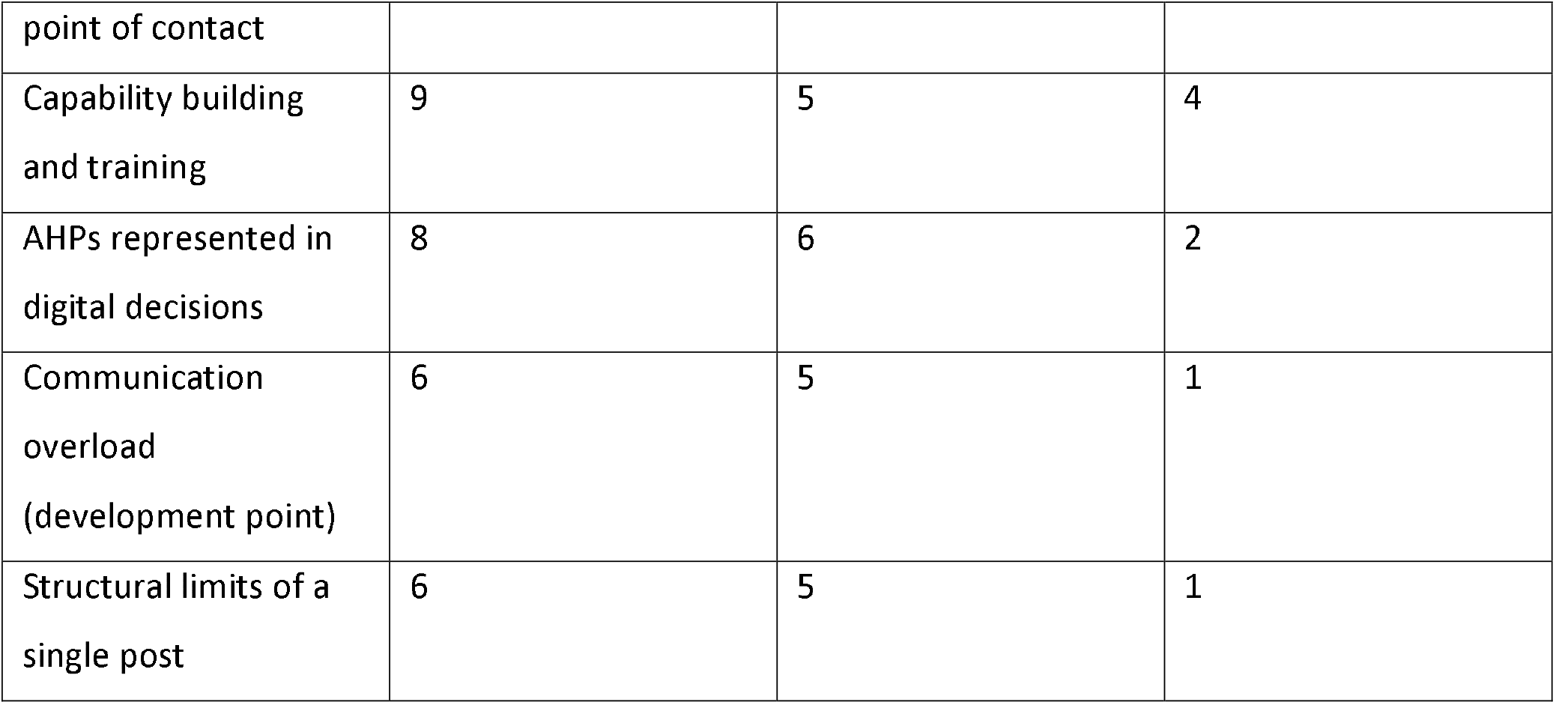
Themes from the coded free text (26 commenters). Every count traces to the respondent-by-theme matrix (Supplementary File, S3).

The data-and-reporting theme ran both ways (14/26): some services described new dashboards and bespoke reports while others still could not get usable, consistent data, locating the cause upstream in analyst capacity and the reporting pipeline. Two development themes complete the picture: communication overload (6/26) and the structural limits of a single post (6/26). Asked what would change if the post were removed, clinical respondents described losing the route by which their issues surface at all: “issues relevant to AHP patients would not be raised” (clinical respondent).

The removal question was answered by all 27 respondents: asked what would stop and what would carry on if the post were removed, they distinguished the day-to-day running of the EHR, which they expected to continue unaffected, from the surfacing, coordination and representation work they expected to stop or slow. That distinction repeats the rating pattern, endorsement of the role’s added layer rather than of system operation, and it is a second sign of discriminating rather than blanket judgement.

A tenth candidate, the value of connecting the service to peer organisations using the same record, surfaced only in the final two responses; it was flagged in the analytic memo and is held as provisional rather than counted, pending replication (Supplementary File, S3).

The praise discriminates. The same respondents who endorse the role name unresolved problems in the next line, and the criticisms are specific rather than diffuse, which strengthens the credibility of the positive ratings from a non-anonymous sample.

### A working framework

Read together, the two strands form a working framework of AHPIO practice and perceived value, drawn as a mechanism rather than a list (Figure 1). One enabling condition, a named, trusted, responsive point of contact, feeds four endorsed functions: brokerage and routing, clinical-digital translation with capability building, representation in digital decisions, and outcome response. Their shared mechanism is connective work, clinical need joined to digital capability, and it produces the near-unanimous perceived value alongside the countable episodic wins respondents describe. A reinforcing loop is proposed from these accounts, for longitudinal testing rather than as a demonstrated dynamic: responsiveness is experienced, trust renews, and issues keep being raised, which is what keeps the role fed; its proposed inverse, ideas met with silence, is how such roles would starve. One pathway is part-built and drawn as blocked: self-service data and reporting, the lone rating shortfall, whose obstruction sits upstream of the post in analyst capacity and the reporting pipeline, so no single role can clear it alone. Operating constraints, the limits of one post, communication load and untested key-person dependence, moderate every pathway.

**Figure 1.**
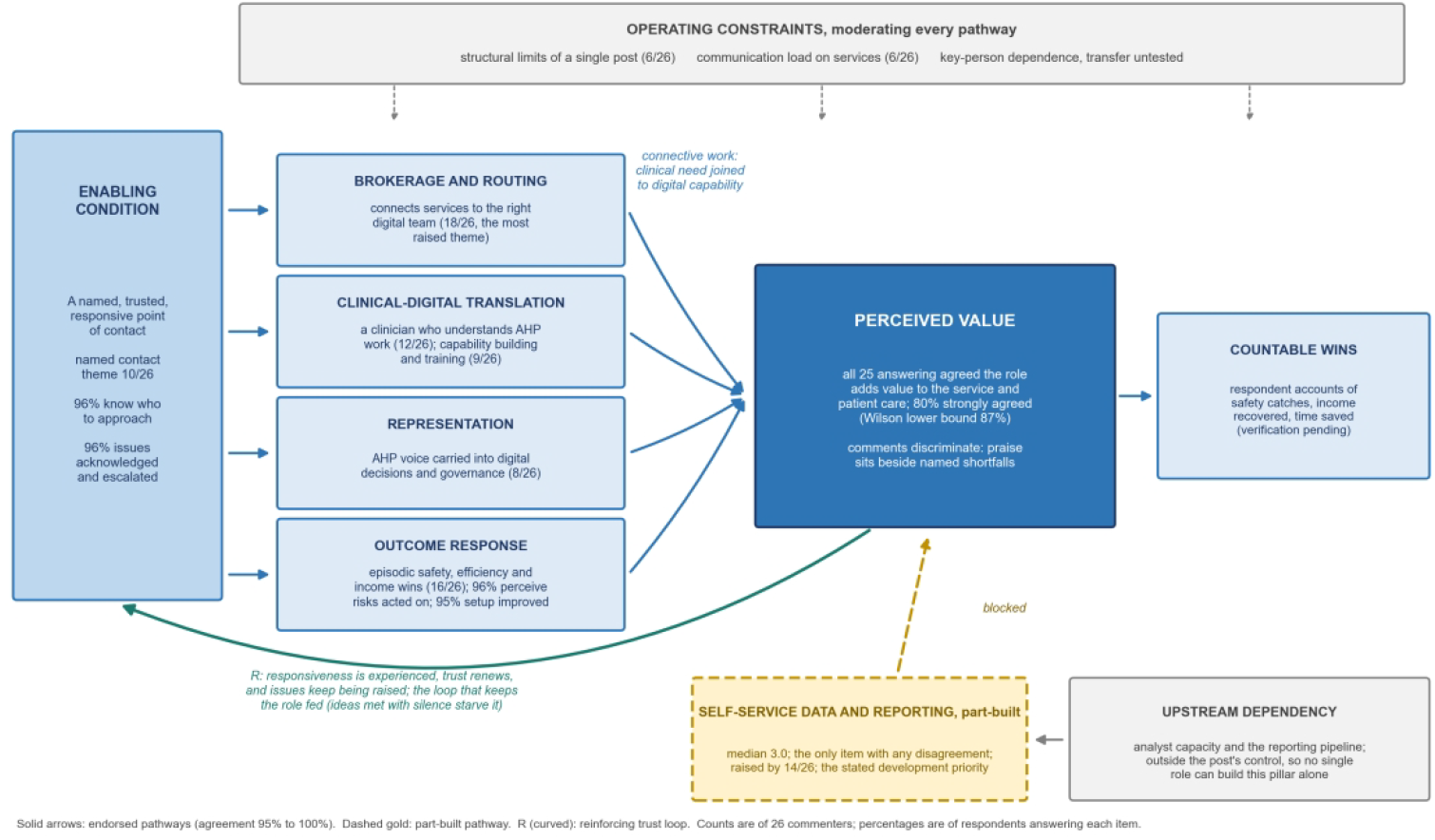
AHPIO practice and perceived value: a working framework drawn as a mechanism. A named, trusted, responsive contact enables four endorsed functions whose connective work produces perceived value and countable wins; experienced responsiveness renews trust in a proposed reinforcing loop (R, hypothesised from the accounts) that keeps issues flowing to the role; the data-and-reporting pathway is part-built and blocked upstream of the post; operating constraints moderate every pathway. Percentages are of respondents answering each item; counts are of 26 commenters.

**Figure 2.**
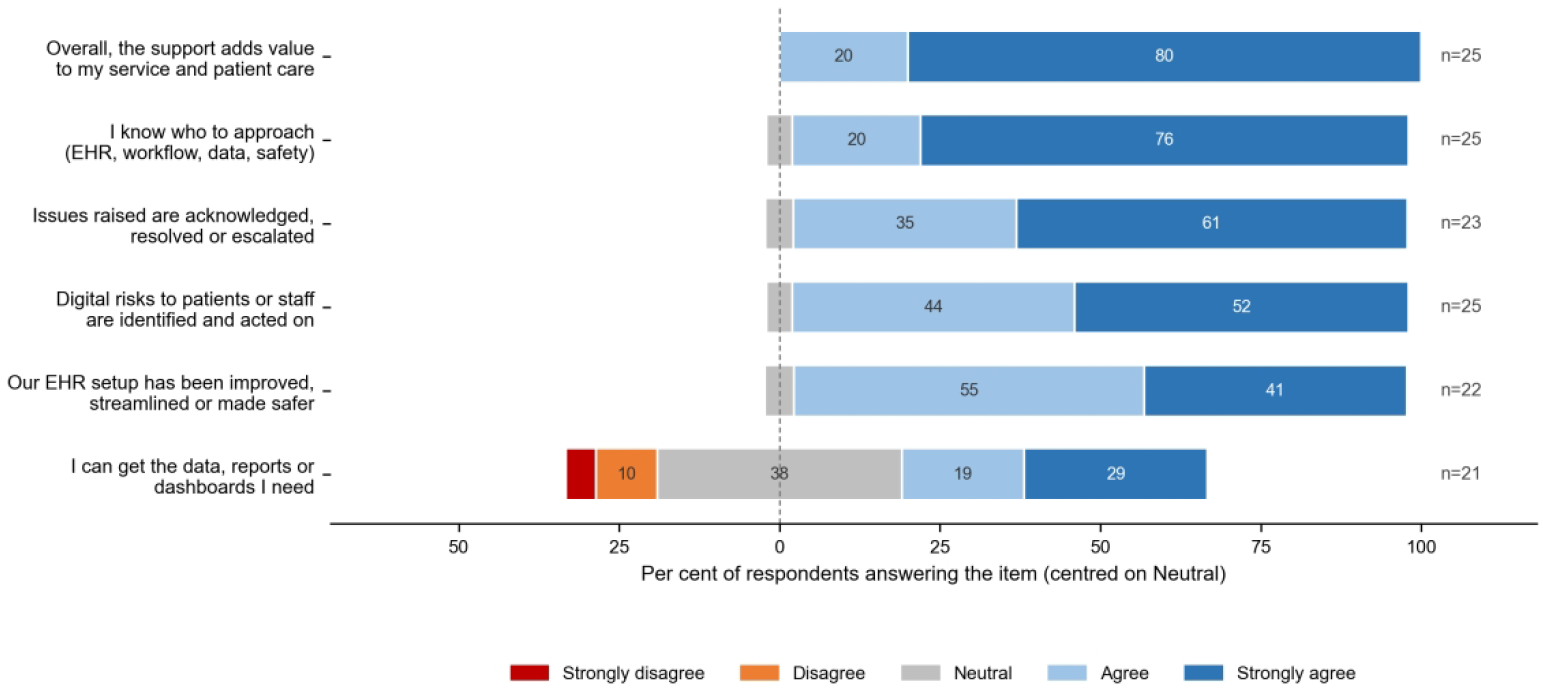
Stakeholder ratings by item, centred on Neutral (per cent of those answering each item; n shown per row). Segment percentages are rounded independently and may not sum exactly to the item totals in Table 2.

## Discussion

Six months into a new dedicated AHP informatics post, stakeholders rated its value at a ceiling on five of six dimensions and located that value precisely: not in operating a system that already runs without the post, but in brokerage, translation, representation and response, resting on a named trusted contact. The one shortfall they identified, self-service access to data, reports and dashboards, is the same capability the free text names as the gateway to a data-led way of working, and it sits upstream of the role in analyst capacity and reporting infrastructure. The evaluation therefore returns a clear development priority alongside its endorsement, and the discriminating pattern of the comments, praise beside named unsolved problems, argues the endorsement is more than courtesy toward a known colleague.

The framework gives the finding a transferable shape, and its most testable element is the proposed reinforcing loop: a role of this kind survives on responsiveness, because responsiveness is what keeps stakeholders raising the issues that constitute its work. Nationally, AHP digital leadership is the weakest assessed domain and formal AHP informatics leads are rare^2^; this evaluation suggests what such a post does in practice while the literature on refinement after go-live explains why the brokerage and translation pillars matter: sustained EHR value comes from clinically informed iteration, which requires someone who holds both the clinical workflow and the system.^5 6^ The pillars also explain why the role’s value is easy to miss: brokered, demand-modifying work leaves few countable traces, so a stakeholder-perceived-value method of exactly this kind, small, named and fully auditable, may be the appropriate first evaluation for emerging informatics roles, ahead of activity metrics that cannot see the work.

### Comparison with existing evidence

Direct comparators are scarce, which is part of the finding. National assessment places AHP digital leadership at the bottom of the maturity profile and formal AHP informatics leads in a minority of organisations,^2^ so evaluations of such posts have had few subjects to study. The nearest quantitative signal comes from EHR refinement work in other clinical groups, where informaticist-supported, clinically led iteration improved clinician satisfaction and reduced documentation burden,6 and from national policy argument that clinically held informatics capacity is the mechanism by which records become useful after go-live.5 7 This evaluation adds the stakeholder’s-eye view of what an AHP-specific version of that capacity does, and a first structured description of where its value concentrates.

### Unanswered questions and future work

Four tests would move this evidence on. The instrument needs validation work before any cross-site use: reverse-keyed items, unbundled constructs and item-level content indexing. The episodic outcome accounts, the safety catches, income recoveries and efficiency gains respondents describe, should be verified against organisational records, which would convert perceived value into demonstrated value where the accounts hold. A repeat administration at twelve to eighteen months would test stability, and any change of post-holder would test whether the value belongs to the role or the person, the key-person dependence this design cannot separate. And the framework invites multi-site replication: if the four pillars and the part-built data pillar recur elsewhere, the framework hardens; where they do not, the differences will be informative.

### Strengths and limitations

The sample is purposive and self-selected, the survey non-anonymous and about named support, so ceiling-bound positivity is expected and the results describe these respondents, never the role in the abstract, a future post-holder, or other organisations. Nine of 27 respondents help produce the work they rated, and their ratings are corroborating context, not independent appraisal; the clinical-only analyses are reported for this reason. The instrument was written in-house, is not validated, bundles constructs in two items and keys all items positively. Two items record perceived outcomes, and no outcome was independently verified. The post-holder authored the evaluation and performed the free-text coding alone, with no second coder; the mitigations stated in the Methods apply, the published verbatim corpus and matrix exist precisely so others can check or re-code the material, and every claim is bounded to satisfaction and perceived value. Against these limits stand a defined sampling frame with a high response for its type, intervals on every headline proportion, exploratory inference that survives its own restriction to clinical respondents, a published verbatim corpus and coding matrix, and full computational reproducibility.

### Implications

For organisations creating AHP informatics posts, the framework offers a starting role description: protect the foundation (a named, reachable, responsive contact), staff the four pillars, and invest early in the data pillar, which no single post can build alone. For the wider agenda, the method travels: a small stakeholder survey with a published free-text corpus and coding matrix is a repeatable, low-cost template for making demand-modifying informatics work visible, and for testing whether this framework holds beyond one centre, one record and one post-holder.

## Conclusion

Stakeholders of a dedicated AHP informatics role endorsed its value near-unanimously and described that value in consistent, specific terms: brokerage, translation, representation and outcome response, on a foundation of trusted responsiveness, with self-service data the one part-built pillar. The framework and the evaluation method are offered as templates for the growing number of organisations building AHP informatics capacity.

## Supporting information

Reporting Checklist

## Data Availability

All data produced in the present study are available upon reasonable request to the authors

## Statements

### Contributorship

DDB is the sole researcher: DDB conceived and designed the evaluation, collected the data, performed all quantitative analysis and free-text coding, wrote the manuscript, and is the guarantor.

### Ethics approval

This study is a service evaluation. The University of Winchester Ethics Committee approved the parent MRes project (Developing Digitally-Enabled AHP: A Mixed-Methods Service Evaluation; reference UREC260401_Bayquen.24; 29 April 2026) through its self-declaration route, under which the project was confirmed eligible for approval without full ethical review. The survey reported here was conducted at the host organisation under its registered service evaluation (SE1576); under the Health Research Authority decision framework the work is not research and NHS Research Ethics Committee review was not required. Participation was voluntary and completion of the survey was taken as consent.

### Data availability

The de-identified response export and the analysis build script are available from the corresponding author on reasonable request.

### Patient and public involvement

Patients and the public were not involved in this staff survey.

