## Supplementary material for "Perceived value and stakeholder experience of a dedicated allied health professions informatics role in a specialist cancer centre: a mixed-methods service evaluation survey": Reporting Checklist

**Reporting checklists: STROBE (cross-sectional) and CHERRIES**

Manuscript: Perceived value and stakeholder experience of a dedicated allied health professions informatics role in a specialist cancer centre: a mixed-methods service evaluation survey. Two EQUATOR-listed checklists are completed: STROBE for the cross-sectional design and CHERRIES for the e-survey administration. Page numbers refer to the revised manuscript of 28 July 2026 (page numbers in the footer); S numbers refer to the Supplementary File (S1 to S6).

**STROBE checklist for cross-sectional studies**

| Item | Recommendation (abbreviated) | Where reported (page) |
|---|---|---|
| 1 Title and abstract | Design in title or abstract; informative abstract | Title and structured Abstract, page 1 |
| 2 Background/rationale | Scientific background and rationale | Introduction, paragraphs 1 and 2, pages 1 to 2 |
| 3 Objectives | Specific objectives | Introduction, final paragraph, page 2; Abstract, Objectives, page 1 |
| 4 Study design | Key elements of design | Methods, Design and setting, page 2 |
| 5 Setting | Setting, locations, relevant dates | Methods, Design and setting, page 2; Data collection (26 May to 16 June 2026), page 3 |
| 6 Participants | Eligibility, sources, selection | Methods, Participants, page 2 (purposive census of the post's known stakeholders, enumeration described) |
| 7 Variables | Define all variables | Methods, Instrument, page 3; instrument verbatim in S1 |
| 8 Data sources/measurement | Sources and measurement methods | Methods, Instrument and Data collection, page 3 (closed online form; internal test submission; non-anonymous by design) |
| 9 Bias | Efforts to address bias | Methods, Reflexivity and competing interests, page 4; Discussion, strengths and limitations, page 7 |
| 10 Study size | How size arrived at | Methods, Participants, page 2 (census of 40; no a priori sample size calculation, stated; exploratory inference declared) |
| 11 Quantitative variables | Handling of quantitative variables | Methods, Analysis, page 3 (ordinal treatment; medians lead; means for convenience) |
| 12 Statistical methods | All methods incl. missing data | Methods, Analysis, page 3 (Wilson intervals; Friedman; pre-stated many-to-one Holm post-hoc family, read descriptively; one-sample Wilcoxon; Mann-Whitney; per-item denominators; no imputation; no weighting); S2 |
| 13 Participants (results) | Numbers at each stage | Methods, Data collection, page 3 (28 returned; test record removed) and Results, Respondents, page 4 (27 of |

| | | |
|---|---|---|
| | | about 40, about 68%) |
| 14 Descriptive data | Characteristics of participants | Results, Respondents, page 4; Table 1, page 9; S2 |
| 15 Outcome data | Numbers of outcome events or summary measures | Results, Ratings, page 4; Table 2, page 9; Figure 2 (legend page 10) |
| 16 Main results | Unadjusted estimates with precision | Results, Ratings, page 4 (all point estimates with Wilson 95% CIs); Table 2, page 9 |
| 17 Other analyses | Subgroup and sensitivity analyses | Results, pages 4 to 5 (clinical-only Friedman; clinical-versus-programme Mann-Whitney; one-sample tests with exact figures; alpha diagnostics); S2 |
| 18 Key results | Summary with reference to objectives | Discussion, first paragraph, page 6 |
| 19 Limitations | Sources of bias and imprecision | Discussion, strengths and limitations, page 7 |
| 20 Interpretation | Cautious overall interpretation | Discussion, pages 6 to 7 (claims bounded to perceived value; the reinforcing loop labelled a proposed dynamic) |
| 21 Generalisability | External validity | Discussion, strengths and limitations, page 7 |
| 22 Funding | Source of funding | Statements, Funding, page 7; also title page |

**CHERRIES checklist (full itemisation)**

| CHERRIES item | Where reported (page) / response |
|---|---|
| Describe survey design | Methods, Design and setting and Participants, page 2: closed survey of a purposive census of 40 known stakeholders |
| IRB approval | Statements, Ethics approval, page 7: University of Winchester Ethics Committee self-declaration route (UREC260401_Bayquen.24, 29 April 2026); registered service evaluation (SE1576) at the host organisation; REC review not required |
| Informed consent | Methods, Data collection, page 3 (voluntary; submission taken as consent) and Statements, page 7; optional named-contact field |
| Data protection | Methods, Data collection, page 3: responses held on the organisation's form platform; names and email addresses removed before analysis; also Statements, page 7, and S4 |
| Development and testing | Methods, Instrument, page 3: items written in-house from a local digital-practice inventory; one internal test submission checked the form before launch and was removed from the dataset |
| Open survey versus closed survey | Closed (invitation-only list); Methods, Participants, page 2 |
| Contact mode | Email invitation to the enumerated stakeholder list; page 2 |
| Advertising the survey | None; direct invitation only; page 2 |
| Web/e-mail | Web-based online form; page 3 |
| Context | Single specialist cancer centre; the post's stakeholder network; page 2 |

| | |
|---|---|
| Mandatory/voluntary | Voluntary; Methods, Data collection, page 3 |
| Incentives | None; Methods, Participants, page 2 |
| Time/date | 26 May to 16 June 2026; Methods, Data collection, page 3 |
| Randomisation of items | None; reported in this checklist |
| Adaptive questioning | None; reported in this checklist |
| Number of items | Six rating items, four free-text prompts, four profile items plus an optional named-contact field; Methods, Instrument, page 3, and S1 |
| Number of screens | Small number of form pages; exact count not retained by the platform; reported in this checklist |
| Completeness check | Not forced; item non-response visible and reported per item; Methods, Analysis, page 3 |
| Review step | No dedicated review page; answers remained editable before submission on the platform; reported in this checklist |
| Unique site visitor | Not separable on the platform; invited list is the frame; reported in this checklist |
| View rate | Not captured by the platform; reported in this checklist |
| Participation rate | 27 of 40 invited (68%); Results, Respondents, page 4 |
| Completion rate | All 27 analytic respondents submitted; two skipped most rating items, reported per item; Results, page 4 and Table 2, page 9 |
| Cookies used | No cookies analysis performed; reported in this checklist |
| IP check | Not performed; closed invited list; reported in this checklist |
| Log file analysis | Not performed; reported in this checklist |
| Registration | Invited list served as the register; no duplicates identified; reported in this checklist |
| Handling of incomplete questionnaires | Included; per-item denominators reported; no imputation; Methods, Analysis, page 3 |
| Questionnaires submitted with an atypical timestamp | One 73-second test record identified by timestamp and removed; Methods, Data collection, page 3 |
| Statistical correction | Holm adjustment within the stated post-hoc family; no weighting (purposive census); Methods, Analysis, page 3 |
