## Supplementary Materials for "Perceived value and stakeholder experience of a dedicated allied health professions informatics role in a specialist cancer centre: a mixed-methods service evaluation survey"

**Supplementary File: Perceived value and stakeholder experience of a dedicated AHP informatics role**

**S1. The survey instrument, verbatim**

The instrument is reproduced verbatim as deployed. The deployed wording names the organisation's EHR product; the brand name is retained here because the instrument is quoted, while the manuscript text refers throughout to the EHR.

Profile: Your role. (Operational/professional lead; Digital champion/superuser; Administrative; Analyst; other, free text)

Profile: Your main service/profession (AHP and/or wider).

Profile: Your main site.

Profile: How often have you needed support in the last six months? (At least once weekly; At least once monthly; Once or twice)

Optional: Name and Email Address.

R1. I know who to approach for help with Epic, workflow problems, service data needs, and digital safety in my area.

R2. When I have raised a digital or Epic issue affecting my service, it has been acknowledged and either resolved or escalated appropriately.

R3. The way my service uses and is set up in Epic has been improved, streamlined, or made safer through this support.

R4. Risks to patients or staff arising from how we use digital systems are identified and acted on.

R5. I can get the activity or performance data, reports, or dashboards I need to understand and run my service.

R6. Overall, this dedicated AHP digital support adds value to my service and to patient care.

(R1 to R6 on a five-point scale: Strongly disagree; Disagree; Neutral; Agree; Strongly agree.)

F1. If you think any rating above needs context, please note which ones and why. (optional)

F2. Think of a specific occasion in the last six months when this support made a difference, or a time you needed it and it was missing. What happened, and what was the effect?

F3. What is working well in the digital and EHR support you receive, and what is not working well or where do you still feel unsupported? Both polar opposites are welcome.

F4. Epic has been live for three years and runs day to day without this post. If this dedicated support were removed: (a) what would stop happening, get slower, become less safe, or go unowned for you; (b) what would carry on unaffected?

**S2. Full quantitative results**

Analytic sample: 28 records returned; one test record (completed in 73 seconds) removed; N=27. Fieldwork 26 May to 16 June 2026. Groups: clinical/operational (n=18: operational and professional leads 10, digital champions and superusers 5, administrative leads 3; services included occupational therapy 4, physiotherapy 3, psychological services 3, speech and language therapy 2, dietetics 1, lymphoedema 1, diagnostic radiography 1, therapeutic radiography 1, plus AHP lead and administrative team lead) and EHR programme (n=9: analysts 8, training 1).

Item distributions and statistics: as manuscript Table 2, computed from the raw export and re-derived independently with no discrepancies. Exploratory inference: Friedman across the six items chi-squared 36.3, df 5, $p<0.001$, Kendall's W 0.36 (20 complete cases); the pattern survives among clinical staff alone ($p<0.001$, 17 complete cases). Holm-adjusted post-hoc Wilcoxon signed-rank comparisons, data access versus each other item: versus overall value $p=0.001$ (Holm $p=0.005$, rank-biserial -1.00, 13 pairs); versus knowing who to approach $p=0.003$ (Holm 0.010, -0.83, 16); versus issue resolution $p=0.005$ (Holm 0.011, -0.77, 16); versus risk response $p=0.003$ (Holm 0.010, -0.82, 16); versus setup improvement $p=0.010$ (Holm 0.011, -0.73, 15). One-sample Wilcoxon versus the midpoint: five items at ceiling (all $p<0.001$, rank-biserial +1.00); data access marginally above neutral ($p=0.040$, rank-biserial +0.63). Zero-difference pairs are excluded under the Wilcoxon signed-rank procedure, so pair counts refer to non-zero pairs and rank-biserial effect sizes are computed on those pairs only. Mann-Whitney clinical versus programme: no detected differences (for example overall value $p=0.694$; data access $p=0.483$), underpowered, and absence of difference is not equivalence. Internal consistency, indicative only: six-item Cronbach's alpha 0.71 (20 complete cases); four operational items 0.83; dropping data access yields a five-item alpha of 0.80. Ceiling effects on five items restrict variance and make these statistics unstable at this size.

**S3. Thematic coding summary**

Free text from 26 commenters was coded by the author, the sole researcher, to nine themes; the analytic memo in section S6 records the coding decisions and alternative readings, and every manuscript count traces to the matrix below. One further candidate (the value of connecting the service to other organisations using the same EHR) surfaced only in the two most recent responses, was flagged during coding, recorded in the analytic memo, and is held as provisional, not counted. Theme counts with group splits are the manuscript's Table 3. The full respondent-by-theme matrix is reproduced below; de-identified verbatim responses are reproduced in section S5.

Respondent-by-theme matrix (26 commenters; a filled cell marks the theme present in that respondent's answers). T1 clinician-understands; T2 brokerage; T3 named contact; T4 concrete outcomes; T5 representation; T6 data and reporting; T7 structural limits; T8 communication; T9 capability.

| ID | Group | T1 | T2 | T3 | T4 | T5 | T6 | T7 | T8 | T9 |
|---|---|---|---|---|---|---|---|---|---|---|
| 2 | Clinical | | X | | X | | | | | X |
| 3 | Clinical | X | | | X | | X | X | | |
| 4 | Clinical | X | X | | X | | | | | |
| 5 | Clinical | | | X | X | | X | | X | |
| 6 | Clinical | X | X | | X | | X | X | | X |
| 7 | Clinical | | X | | | | X | | X | |
| 8 | Clinical | X | X | X | X | | X | | | X |
| 9 | Clinical | X | | X | | X | X | X | | |
| 10 | Clinical | X | X | X | X | X | | | X | |
| 11 | Clinical | | X | X | X | | | | | |
| 12 | Programme | | X | | X | | | | | |
| 13 | Clinical | | X | | X | | X | | X | |
| 14 | Clinical | | X | | | X | X | X | | |
| 15 | Programme | X | X | | X | | | | X | |
| 16 | Clinical | | X | | | X | X | | X | X |
| 17 | Programme | | X | X | | | X | | | X |
| 18 | Clinical | X | X | X | X | | X | | | |
| 19 | Clinical | | | | X | X | X | X | | |
| 20 | Programme | | X | | | X | | | | |
| 21 | Programme | | X | | | | | | | X |
| 23 | Clinical | X | X | X | X | X | | | | X |
| 24 | Programme | X | | | | | | | | X |
| 25 | Clinical | | | X | X | | X | | | |
| 26 | Programme | X | X | | X | | | | | |
| 27 | Programme | | | | | X | | X | | X |
| 28 | Clinical | X | | X | | | X | | | |

**S4. Condensed CHERRIES summary**

The compulsory reporting-guideline upload is the separate checklist document (STROBE cross-sectional and full CHERRIES, item by item with page numbers); this section is the condensed summary.

| CHERRIES item | This survey |
|---|---|
| Design | Cross-sectional closed online survey of a purposive census of a post's 40 known stakeholders. |
| Ethics and consent | University of Winchester Ethics Committee self-declaration route (UREC260401_Bayquen.24, 29 April 2026); registered service evaluation (SE1576) at the host organisation; REC review not required. Participation voluntary; submission taken as consent; a named-contact field was optional. |
| Data protection | Responses held on the organisation's form platform; names and email addresses removed before analysis. |

| | |
|---|---|
| Development and testing | Items written in-house from a local digital-practice inventory; one internal test submission checked the form and was removed from the dataset. |
| Open versus closed; contact mode | Closed; invitation by email to a defined stakeholder list; no advertising. |
| Administration | Online form, 26 May to 16 June 2026; completeness not forced; no dedicated review page, answers editable before submission; no incentives. |
| Items | Six rating items, four free-text prompts, four profile items and an optional named-contact field (verbatim in S1). |
| Response | 27 of 40 invited (68%); view rate not captured by the platform; no duplicates identified; one test record removed on its timestamp. |
| Analysis | Only the test record excluded; item non-response reported per item; no imputation; Holm adjustment within the pre-stated post-hoc family; no weighting. |

**S5. De-identified verbatim free-text responses**

Responses are reproduced verbatim by question, attributed by respondent number and group only (clinical or programme), because several services have a single respondent. The deployed wording names the organisation's EHR product and a legacy imaging system; these names are retained because the material is quoted, while the organisation's own programme name is redacted in brackets. Square brackets mark redactions made at source or here.

**F1. Context for any rating (optional)**

P3 (clinical). I feel that the role needs more support, although, I fully appreciate NHS financial constraints. For me to get the reports that I want, this role is helping enormously but I guess with all the digital work and limited workforce impacts on fully resolving such issues

P4 (clinical). I do not raise many issues to Epic but delegate to my team.
The AHP IO has been particularly helpful assisting in a complaint and successfully navigating EPIC.

P5 (clinical). [AHP IO] has been such an asset to the team. We (Therapies) couldn't function without him in his role. Issues have been resolved quickly and the changes he has supported us to make have improved income generating workflows (e.g. PP billing) and efficiencies.

P6 (clinical). Not all of our data is available in Slicer Dicer  -the analysts and IO are aware and are working to sort this out. Some of the reports we need are complex and need an analyst to build them

P9 (clinical). AHP support for epic I feel has improved over the past 5-6 months with good access to support and feedback on actions and incidents. It would be good for me to get improved data and analytics for my service but I haven't raised this as a current concern so don't expect this to be available necessarily. Epic has been improved to some extent off the back of concerns an issues raised but within limitations of the system and the trust needs.

P10 (clinical). Since coming into the role, there has been an increased amount of time spent supporting AHPs utilise and optimise EPIC. This role has definitely improved our workflows.

P13 (clinical). I still cant get the data I want out of EPIC but I understand that this is a process more widely needed across therapies

P14 (clinical). I use radiology system lead for a the majority of immediate or long term data / activity required for B cases, dashboards, performance as we use 3 systems ( Soliton/ EPIC and PACS)

P17 (programme). As an Epic trainer, I really appreciate the support of [Nursing IO] and [AHP IO] when I have questions about operational issues. So I do feel very supported by the NIOs.

P18 (clinical). Activity data - we have bespoke reports created for us which is nice. does epic provide dashboards ?

P19 (clinical). Capacity in the system seems to be the biggest constraint ie the analysts can only take on so much and understandably work has to be prioritised. It has been very useful having a role where the clinical and the digital contexts are understood.

P20 (programme). I am a Configuration Manager within the [the EHR programme] team and have found it very helpful having an AHPIO (in addition to the NIO team) where support about requests/projects involving end users and workflows we are less familiar is available. Frequency of contact is dependant on activity with operational teams who are not our main end users so may increase if there is a project or ticket involving my team.

P24 (programme). I am an Epic Trainer who also support our helpdesk so some of the above do not match my role

P26 (programme). N/A to some answers as I am analyst and not a service user

P28 (clinical). Reporting remains a problem - especially being able to demonstrate the effectiveness of the service.

F2. A specific occasion when the support made a difference, or was missing

P2 (clinical). I have been able to resolve issues quickly with support and had a workflow streamlined which made seeing patients much more efficient

P3 (clinical). It is helping me to create a performance and productivity dashboard. In addition, [AHP IO] is also helping me create a rehabilitation summary and dashboard, this will support safe, and effective patient flow and discharge.

P4 (clinical). As mentioned the assistance I received with a complaint ensuring I had not missed any vital entries on EPIC.

In addition we have been sorting out PP payments to Therapies which has been essential as Lymphoedema were not being charged- massive oversight.

P5 (clinical). Our lymphoedema workflows were not adequately set up for PP billing. This was identified and quickly rectified. This ensures accurate and appropriate billing for services provided.

P6 (clinical). As the result of an Incident it was identified that the referral process for RIG tube insertion needs to be changed to add a dietitian as a co-sign to prevent inappropriate tubes being inserted. This has been picked up by our Epic team and the adjustments required are being developed and our AHP IO is supporting. This is an example of how the changes will make a significant improvement to patient safety.

P7 (clinical). Digital walkarounds have been really useful for queries to be tackled in a straight forward manner.

P8 (clinical). I altered [AHP IO] when we noticed after an upgrade that OP visits were automatically signed off even if work not finished. He raised this with analysts.

P9 (clinical). Appointments being edited by administrators external to department. issue was raised and actioned as much as possible by AHP IO alongside admin managers.

P10 (clinical). The AHP IO supported the OT team with the optimisation of their outpatient navigator. It had been a difficult process due to the OTs not having been through optimisation previously. The AHPIO was able to explain the process, supported with making smart phrases and advise on the implications of certain flow sheets. As the AHP IO understands the role of AHPs he was able to provide profession specific help which was really useful. The AHP IO is supporting was reducing documentation time but supporting with flow sheet advice and creation.

P11 (clinical). Cancelled appointments by other services affecting APSS workflows - issue raised and immediate contact made to support team and service

P12 (programme). Support provided in moving forward a project to digitise mortuary workflows. Assistance in organising and relating information to the SME group by the AHP IO was extremely helpful in moving the project over the line.

P13 (clinical). We have had our full navigator optimised which has been really useful and will be easier to document for patients. It has been key to have the IO involved in this to help move it forward.

P14 (clinical). we have optimisation meetings with representation from relevant leads - where issues are rated / scored and plans made and support advice required

P15 (programme). A recent example was the work around the Contrast workflow transition from Soliton to Epic. The AHP EHR support team helped us coordinate with the right end users, understand the clinical requirements, and validate the workflow end-to-end. Their involvement meant I could configure the system correctly, avoiding delays and rework. This support made the process smoother, safer, and more efficient for both clinicians and patients.

P16 (clinical). I have been asked to keep on top of stats in my team and initially didn't have the dashboard to check OT stats. Now i have.

P17 (programme). [Nursing IO] was extremely helpful when I delivered the thrive training for psychologists. Her input, knowledge and support during the session was invaluable as she answered questions and took the lead in a way I couldn't do. Sorry, I can't think of a concrete example of a question, but I was very glad she was involved.

P18 (clinical). we have had a number of examples. we wanted a bespoke service to document unwell pats in RT- the epic created something very helpful and even suggested a better solution to this.

P19 (clinical). [AHP IO] attended the physical activity strategy meeting and helped us to explore digital options for collecting physical activity data

P20 (programme). Support clearing old tickets advising where still required and who to involve/contact for queries and validation

P21 (programme). Bridging the knowledge gap between Analysts and end users. Users sometimes add more tasks to solve their problems. The Information officers have ben able to look at the scenario and identified the exact needs and ways of resolution. Most of the issues are training related and the Information officers have been able to suggest/point to the appropriate tools needed.

P23 (clinical). Our team is currently transitioning to becoming paperless, and having been shown how to padlock notes as sensitive will be very helpful in this transition.

P24 (programme). I do not think as an Epic trainer I can answer this, as I would be the one giving support via our Helpdesk or training sessions. I get support from all of my colleagues including other Epic teams and they are always extremely helpful and knowledgable.

P25 (clinical). A specific example where this support made a difference was when I needed clarification on which report would be most appropriate to use for creating our performance slides. I contacted [AHP IO], who reviewed my approach and explained that the Time Use report was the most suitable report for this purpose. He also confirmed that the way I was extracting and presenting the data was correct. This reassurance gave me confidence in the accuracy of the information being reported and ensured that the performance slides were produced consistently and using the most appropriate data source.

P26 (programme). I'm involved in project to get Speech and Language Therapists using EPIC to record PGD ordering and administering. The IOs have been helpful in coordinating the project as it involves several different apps. I noticed there was a mistake in the PGD documentation and [AHP IO] organised for this to be fixed by getting in contact with the right people.

P27 (programme). AHPIO was able to drive SMEs to make decisions within established deadlines to ensure projects moved forward.

P28 (clinical). Meeting other Trusts and learning about the way their admin processes work has been invaluable and having the support of the Information Officer to help drive some of these changes is really useful.

Recording non-patient consultations (for family referrals). This has been an ongoing issue since EPIC launch. I feel we are making more headway with this but what is frustrating is more people finding out about this when we are about to implement and more queries being raised. Senior EPIC personnel sat in on initial meetings so i'm unclear as to why this continued to be a problem.

F3. What is working well, and what is not working well or still unsupported

P2 (clinical). More training would help and then time to consolidate learning - it is impossible to apply learning when trying to run a busy clinic, I would just revert to the old, familiar but more time consuming ways, as I don't have time to apply new learning that is unfamiliar e.g. using Dragon. Despite training, there has been no opportunity to implement this into my daily practice.

P3 (clinical). [AHP IO] is enthusiastic and I feel has contributed enormously to the success of this role. We have got far more than expected from this role.

P4 (clinical). so far so good- all well

P5 (clinical). Positives: visibility, presence, proactivity, Therapies tips session after cross site meeting
I don't feel unsupported at all but I receive a high volume of emails and cannot read/action all in a timely way so less frequent, concise emails would be valued.

P6 (clinical). The escalation of our needs and the feedback is really helpful. This means we know when things will be corrected or improved.
It would be beneficial for the IO to spend some time shadowing us to understand more intimately how we interact with Epic and then to analyse and show us how to make efficiencies, use it better and get clean data from it to support our service and interventions

P7 (clinical). Obtaining data from Epic is still very frustrating as the data seems to be inconsistent when pulled from different sources, and it is really hard to find a data collection structure that works well and reflects what happens on the ground. I feel like the volume of digital email communications is sometimes much, and often I do not understand a lot of it (sometimes seems like it's written in riddles), therefore they go without being addressed, as more pressing things take priority.

P8 (clinical). I feel it is very helpful to have a named person who is willing to support us with Epic issues and who has dedicated time to focus on this.

I would like to have more time to explore information gathering and how to use data. I feel this is probably possible with support but NO TIME at the moment.

P9 (clinical). I feel it is easy to contact a member of the IO team and get a prompt response so I feel supported in this way.
I think it is still challenging to get optimisation requests worked on promptly due to shorter staffing

in analyst teams unfortunately but understand optimisation projects need to be prioritised based on need.

P10 (clinical). Communicates very clearly with the OT team.
There is a lot of tip sheets for EPIC which can take a lot of time to read and understand - but not sure if the AHP IO can help with this!

P11 (clinical). Approachability of the support has been hugely appreciated in building relationships to resolve issues going forward

P12 (programme). Increased communication regarding AHP needs. Expedited access to appropriate SMEs from the relevant areas to maintain project movement.

P13 (clinical). I find the current process responsive. Having to go to multiple committees to use questionnaires we already use is time consuming.

P14 (clinical). we feel supported , but we work within 3 systems , so there is a balancing act to ensure don't ever make decisions that effect another system/ modality / process  in the other systems / modalities further down the line

P15 (programme). What works well is the ability to reach the correct end users quickly. This makes it easier to understand issues clearly and deliver accurate configuration changes. Their knowledge of the services and their relationships with clinical teams significantly improves communication and speeds up problem resolution.

Areas that could improve include having more structured feedback loops after changes go live, and clearer ownership boundaries between different digital teams. Sometimes it is not obvious which team should pick up certain issues, which can slow things down.

P16 (clinical). Wednesday walk arounds are useful but i do generally forget they are happening.  Is there a system where we can log a request so we know the support person will come around or is it best to email them directly?

P17 (programme). The support I receive from [AHP IO] and [Nursing IO] is fabulous and they are quick to respond to any queries which may arise on the Help Desk or from training. I also think they provide a vital role in supporting therapists in their on-going use of Epic, since induction training is only the very beginning of a therapist's Epic learning. They provide context which is sometime hard to create in the classroom.

P18 (clinical). as soon as we raise a ticket it is actioned immediately

P19 (clinical). I don't feel unsupported but am aware requests have to be prioritised and so solutions can take time. For example we have been waiting a long time for an automated route to be established for pre-surgical breast patients to be booked into the pre-assessment clinics.

P20 (programme). Great to have another IO to advise/escalate/identify appropriate operational contacts when addressing [the EHR programme] tickets/requests/projects

P21 (programme). The SNOW ticket logs works well as I can always check to see the progress of a query.

P23 (clinical). We had been lacking support for a long time (1 year+) but now we have a dedicated and named individual to support us, we have regular check-ins, questions are taken to the relevant person and answered.

P24 (programme). I always offer extra support post training by ways of 1-1's, virtual meetings etc and also supply our support number and email which I hope helps all Epic users.

P25 (clinical). What is working well is the excellent support we receive from [AHP IO]. He consistently goes above and beyond to help resolve queries and provide clear guidance, such as advising us on the correct process for reporting a patient death in Epic.

Overall, I have always felt well supported and cannot think of any occasions where I did not receive the help I needed.

P26 (programme). What works for me is the communication between the IOs and the end users, they know the right people to contact and act quickly, this allows me to focus on the configuration/build that I need to do.
I don't feel unsupported

P27 (programme). I think it's good that each group of healthcare workers now has representation. Unfortunately there are too many areas requiring support and we're unable to cover everything as quick as we wished we could.

P28 (clinical). Having a 'go to' support is really helpful to keep on track with optimisation requests etc as sometimes it can feel like these fall into a bit of a black hole.  As highlighted above, the meeting with other Trusts on how they use EPIC has been invaluable and I'm confident with the support of the Information Officer we will be able to implement better ways of working.

F4. If this dedicated support were removed, what would change

P2 (clinical). There is still a lot of support that is needed to make our clinical work more efficient so without this support we will not be able to meet the organisational objectives.

P3 (clinical). Removal of this role would impact greatly on delivery of rehabilitation services. It would certainly increase the time of our clinical staff dealing with Epic issues, and thus limit clinical capacity for service delivery. It would impact on patient flow and discharge. I feel might also contribute to increased staff turnover

P4 (clinical). Issues would not be escalated in a timely manner. If a non AHP were in the role they would not understand our needs. It makes such a difference to speak with a clinician.

P5 (clinical). Since [AHP IO] has been in post, we have been able to make rapid improvements within an ever evolving system. This has improved efficiencies, productivity and income generation. There is so much more to do and this will be ongoing given the nature of healthcare. We learn such a lot each week and [AHP IO] supports us to utilise our system - both from a notes perspective and a reporting perspective.

P6 (clinical). To be honest I think it's a bit too soon for me to be able to state what would be lost if this post were to go. I think that everything would get slower but some improvements would still happen because of the way that the processes happen and that as a team we tend to request and drive change. I do think that what would be lost is the ability to have a data driven service because an AHP IO understands better the way we function and what we need from Epic

P7 (clinical). We would lose dedicated support and championing for making Epic work better for the team.

P8 (clinical). Without the dedicated support we would have to spend more time explaining the details of our role and specifics for therapies. This is likely to result in repeated requests or further updates needed by the IT team if they do not understand what we are asking for. This is a two way thing as we are probably not good at IT-speak!

P9 (clinical). A) response times to issues and concerns would likely get slower. Having dedicated support for AHP's enables someone who understands the service to appropriately action concerns. B) day to day concerns user errors / questions could be absorbed by superusers. Datix/incidents may be more difficult to absorb by managers linking with correct epic team member. National Epic meetings have been attempted to be absorbed by superusers but this also feels unrealistic to be involved at a good level e.g. being involved in guidelines/competencies being developed, being able to contribute at an operational level to meeting discussion so I feel needs dedicated support

P10 (clinical). Prior to the AHP IO we had not made any steps forward with optimisating EPIC within OT work flow, therefore feel this would have been missed without the AHP IO. We previously were not involved in any updates or new projects within the whole hospital, whereas now I feel we are being included.

P11 (clinical). There are always changes being looked at in Epic and issues arising that services need support with to improve the quality of patient pathways. Without this support services will lose a significant resource available to them to support and implement changes quickly and effectively. The service would be unable to manage the coordination of the support needed to implement such changes and improvements.

P12 (programme). This service has been invaluable in providing connection and communication with all AHP services and the digital team.

P13 (clinical). It would take us a lot longer to do things.

P14 (clinical). its good to have a lead who has oversight into other areas/ meetings with regards to EPIC optimisation lessons learnt etc , but needs constant link with our systems lead re the above conflicts

P15 (programme). If this dedicated support was to be removed I feel the impact would be slower access to end users, more configuration errors, weaker coordination with clinical teams, and increased risk of workflow or safety issues.

Routine configuration or basic triage could shift to central teams and be absorbed, but the specialist AHP knowledge and relationships would be difficult to replace.

P16 (clinical). a) potentially issues relevant to AHP patients would not be raised so services/ methods wouldn't be reviewed or improved
b) potentially being the support person for the team , such as weekly walk around . I know that's what the digital champions are meant to be for, but perhaps increasing their confidence to do would help this .

P17 (programme). I think it's vital to have someone I as a trainer can contact with questions. I also think it's vital for therapists to have on-going support in developing their knowledge and use of Epic. On-going support is vital to having quality data in Epic. For example, [Nursing IO] has been instrumental in encouraging therapists to create a telephone encounter for indirect work they do for patients e.g. phoning suppliers. I now explain in training that telephone encounters can be used to assess how much time is being spent on activities for patients which a documentation only encounter can't assess.

P18 (clinical). a) it would potentially get slower. having the IO is good to have a point of contact especially for AHPs as we are such a bespoke team. I have emailed the IO and have had immediate responses. When EPIc had downtime the IO support was really helpful.
b) the epic team would need to absorb any simple questions, with the volume of incidents being increased. The service would need additional support for some EPIC related changes and training

P19 (clinical). We can work on efficiency measures with the support of this post for example filling cancellations from our OP wait list; whilst this could be completed by other posts I suspect it is less likely to be prioritised without and AHP voice. As managers we don't have the right connections into the digital world to influence effectively.

P20 (programme). More pressure on NIO team to help advise/support as before position existed. Assumably less satisfaction from non-medical/nursing/pharmacy operational teams particularly for projects or more significant changes as no representative at DHR fora

P21 (programme). Requests will get lost or forgotten

P23 (clinical). a) Our services' specific needs would not be considered or addressed, and we therefore may not be using EPIC in the best way possible.
b) The EPIC team would have to support us instead.

P24 (programme). If this support were removed, I guess it would make me redundant!!! It would definitely be a worry when policies and needs change as no one would be available to update the system and educate users. I think staff can use the Tipsheets better but again, these would be written by Epic staff so needs constant maintenance.

P25 (clinical). If this dedicated support were removed, resolving Epic and reporting queries would likely become slower, and staff would have less access to expert guidance on data quality. This could affect efficiency and confidence in using the system correctly.

P26 (programme). a) I think it would take more time for projects to be completed
b) Providing more support/communication with end users - although this wouldn't be ideal

P27 (programme). a) AHPs would likely feel a little neglected as Trust tends to focus on acute areas (nursing and doctors, SACT); b) Teams could have their own digital information leads/champions reviewing requests and filling in forms for new build etc

P28 (clinical). There would be no person monitoring our optimisation requests. This has been invaluable especially that this person can sit in and observe processes to get a greater understanding of our requests. It is often very difficult to put what we would like on to a paper requests.

What continues to be a bug bear is the amount of appointments and referrals that are able to be cancelled by people from outside of our department. I have highlighted this since go live and there have been times where I feel we have got somewhere and then we are back to square one. It has never felt like this has been taken seriously before and this is a risk to patients and their care and treatment by Psychological Services.

**S6. Analytic memo (single coder)**

**Provenance.** Compiled on 28 July 2026 from the study record and the internal report of 16 June 2026; the decisions recorded here were made during the analysis. It is reproduced because the free-text coding was performed by the author alone, so this memo, the verbatim corpus (S5) and the matrix (S3) together carry the audit function a second coder would otherwise serve.

**1. Position.** I hold the post being rated. The risks recorded at the outset were hearing praise as vindication and hearing criticism as user failing. The decisions below were made against those risks, and criticisms are reported unsoftened.

**2. Coding approach.** Codes were kept close to respondents' own words, and the nine theme labels are working phrases drawn from the responses. Each commenter's answers were assigned to every theme they raised; counts are respondent-level, so a respondent contributes once to a theme however often they raise it. The matrix in S3 is the complete record of assignments, and the verbatim corpus in S5 is the complete source.

**3. Decisions recorded.** First, the tenth candidate (the value of connecting the service to peer organisations on the same record) surfaced only in the final two responses; it is held as provisional rather than counted because it arrived too late to be tested against earlier respondents, who were not asked about it. Second, programme-group comments are treated as corroborating context rather than independent appraisal, because that group co-produces the work it rates. Third, accounts of safety, income and efficiency outcomes are coded as respondent accounts, never as verified outcomes. Fourth, the data-and-reporting theme is coded in both directions, wins and gaps, rather than forced to a single valence.

**4. Alternative readings held.** The ceiling could reflect courtesy toward a named colleague rather than value; the discriminating pattern (praise beside named, specific shortfalls, and one item drawing open disagreement) argues against pure courtesy, and the claims are bounded to perceived value regardless. The removal question could invite loss-averse inflation; in the event respondents were specific about what would carry on unaffected, which is the opposite of blanket loss talk.
